# Covid-19 Pandemic- Pits and falls of major states of India

**DOI:** 10.1101/2020.06.18.20134486

**Authors:** Abhinesh Singh, Samriddhi S Gupte

**Affiliations:** Research Associate, Tata Institute of Social Sciences, Mumbai; International Institute for Population Sciences

## Abstract

Covid-19, just like SARS and MERS before it, is a disease caused by corona virus and can lead to severe respiratory diseases in humans. With the outbreak of novel corona virus, WHO on 30^th^ January 2020 declared it a Public Health Emergency and further on 11^th^ March 2020, Covid-19 disease was declared a pandemic. India in the initial stages of the pandemic dealt with it in a very effective manner. With timely implementation of lockdown, India was able to contain the spread of Covid-19 to some extent. However with the recently announced Unlock 1.0, the SARS CoV-2 is expected to spread. This study aims to track and analyze the Covid-19 situation in major states that constitute of 70 percent of the total cases. Thus the states selected for the study are: Maharashtra, Delhi, Tamil Nadu, Gujarat, Uttar Pradesh and Rajasthan. These are the states which had more than ten thousand Covid-19 patients as/on June 10^th^ 2020. The analysis period is from March 25^th^ to June 10^th^ and the data source is India’s Covid-19 tracker. To assess the previous and current Covid-19 situations in these states indicators such as Active rates, Recovery rate, Case fatality rate, Test positivity rate, tests per million, cases per million, test per confirmed case has been used. The study finds that although the absolute number of active cases may be rising, however it is showing a decreasing trend with an increase in recovery rates. With increasing number of Covid-19 cases, testing also has increased however not in the similar proportion and thus by developed nation standard we are lagging. With increasing TPR and cases per million, Delhi is well on its way to surpass even Mumbai which till now has proven to be worst hit in this pandemic. An interesting finding is that of test per confirmed case which shows that every 6^th^ person in Maharashtra and every 8^th^ in Delhi is showing positive result of Covid-19 test. Given such an increase and unlocked India, Delhi might soon enter into the third stage of community transmission where source of 50 percent or more cases would be unknown. There has been an increase in the Covid-19 related health infrastructure with the public-private partnership which involved both private hospitals and lab joining hands to battle Covid-19, however, affordability still remains an issue. If experts are to be believed, pandemic isn’t over because we’ve unlocked. The worst is yet to come as Covid-19 is predicted to peak in mid-July to August in India. Thus, it’d be advisable to not venture out unnecessarily just because restrictions have been lifted. Also, following the guidelines-hand-washing, avoiding public gathering, social distancing and covering nose and mouth has now become imperative.

## Introduction

Covid-19, like its ancestors SARS and MERS is caused by a zoonotic pathogen by the name Corona Virus and can cause severe respiratory diseases in humans ^[1][2]^. World Health Organization on January 30^th^ 2020 declared Novel Coronavirus as Public Health Emergency of International concern and on March 11^th^ 2020 Covid-19 disease was stated as pandemic based on its spreads severity ^[3]^. A peculiar aspect of Covid-19 disease is that its transmission can be symptomatic, mild symptomatic, or even asymptomatic. A study conducted by Indian Council of Medical Research said that out of total active cases till April 30^th^, 28 percent were asymptomatic and it was also observed that such patients were younger and without comorbidities ^[4]^. The said study also evaluated Covid-19 patients by age and stated that the attack rate per million was highest among 50-69 years old and lowest among children below 10 years old. Gender wise attack rate was higher among men compared to women ^[5]^. India has 287157 confirmed Covid-19 cases out of which 48 percent patients constitute the active cases and 49 percent patients have recovered as on June 10^th^ 2020 ^[6]^. In order to check the spread of CoV-2 at an early stage, a nationwide lockdown was implemented in four phases from March 25^th^ to May 31^st^ 2020. Many studies asserted that without lockdown, cases would have tripled or quadrupled in comparison to today’s figure ^[7]^. On April 11^th^ the Joint Secretary of Health Ministry, Lav Agarwal, presented a graph which projected CoVid cases had lockdown not been implemented and stated the scenario would have been grave as the statistic of confirmed cases and doubling rate would have sky rocketed ^[8]^. Also, the World Health Organization Emergencies Programme Executive Director, Michael Ryan, on June 5^th^ said that due to lockdown in India the doubling time for Covid-19 spread is about three weeks at present. ^[9]^.

On 1^st^ June 2020, around 9 weeks after the first phase of lockdown, India announced unlock 1.0. But the question remains whether lifting the lockdown is a good idea. In accordance to the World Health Organization recommendations a nation could unlock only if and when daily positivity rates fall below 5 percent and remain so for a period of 14 days. However, India’s daily positivity rates was nowhere near 5 percent and stood at 7.7 percent (on June 1st) ^[10]^.

With the increasing spread of corona virus infections, it becomes imperative for the testing to increase in the same or at least similar proportion. Testing per million population serves as an important indicator to compare nations in regard to handling Covid-19 disease. Enough testing ensures and gives the actual figures of Covid-19 patients. India, although, has conducted more than 5 million tests yet it is not adequate given India’s population, the figure for its testing per million population remain less than many other Covid-19 affected countries ^[11]^. A study while analysing the burden of pandemic in India found that Maharashtra having highest number of Covid-19 positive cases is solely responsible for more than one third of cases as on May 17^th^ 2020 followed by Gujarat and Tamil Nadu. Also, higher test positivity rate is another issue in these states. Along with this the study also forecasted that Covid-19 designated hospitals are less in number and that soon all the ventilators will be occupied due to increasing number of patients along with other patients suffering from communicable and non-communicable disease amidst this pandemic ^[12]^. This can turn out to be true in forthcoming days as peak of Covid-19 cases is expected in mid-July. Therefore, this study aims to track and analyze the Covid-19 situations of the major states which share almost 70% burden of the total cases. Thus, the states taken into consideration for the study are the states which had more than 10,000 confirmed Covid-19 patients as/on June 10^th^. The six major states so included are: *Maharashtra, Tamil Nadu, Delhi, Gujarat, Rajasthan, and Uttar Pradesh*.

The Covid-19 related information is taken from the Application Programming Interface (API, http://api.Covid19india.org). It is a data sharing portal that provides the most recently updated information on daily basis. It gives data related to total tests, confirmed cases, active cases, recovered cases, and deaths for each affected state/UT as well as for whole India. The number of Covid-19 designated public and private laboratories doing Covid-19 testing has been taken from Indian Council of Medical Research website as they publishes list of laboratories for each states/UTs. The projected population of India and states/UT for the year 2020 have been taken from Report of the technical group on population projections, 2019 ^[13]^.

The analysis for the study has been done from 25^th^ March 2020 (lockdown 1.0) all the way to 10th June 2020 i.e. ten days after unlock 1.0. For sake of convenience in analysis, this time period is further bifurcated in 10 days interval. In the study, rate of active case is defined as the total number of active cases out of total confirmed cases. Recovery rate is defined as the total number of recovered cases out of total confirmed cases. Case fatality rate is defined as the total number of deaths out of total confirmed cases. Test Positivity rate is defined as the proportion of confirmed cases per 100 people tested. Tests per million population and cases per million population are also calculated. Test per confirmed cases is defined as the number of test done per positive cases.

## Findings

**Table 1** gives the Covid-19 scenario of India and states/UT selected for study since 25^th^ March to June 10^th^ 2020. The number of confirmed cases has increased in this duration for the India as well as states/UT with diminishing rate. The study calculated rate of active cases for the selected time period for India and states/UT and it is decreasing since outbreak of coronavirus and recovery rate is also calculated and it is increasing for India and states/UT. Study also calculated case fatality rates and it shows total number of deaths out of confirmed cases.

**Table 1:** **Confirmed cases, Active cases, Recovered cases, Deceased cases, and their respective rates for India and selected states for selected time period**.

| Country/States/UT | Date | Total confirmed cases | Active cases | Active cases rate | Recovered cases | Recovery rate | Deceased | Case fatality rate |
| --- | --- | --- | --- | --- | --- | --- | --- | --- |
| India | 25<sup>th</sup> March | 657 | 650 | 98.9 | 3 | 0.5 | 1 | 0.15 |
|  | 10<sup>th</sup> April | 7599 | 6561 | 86.3 | 786 | 10.3 | 249 | 3.28 |
|  | 20<sup>th</sup> April | 18544 | 14675 | 79.1 | 3273 | 17.6 | 593 | 3.20 |
|  | 30<sup>th</sup> April | 34867 | 24650 | 70.7 | 9059 | 26.0 | 1154 | 3.31 |
|  | 10<sup>th</sup> May | 67177 | 49389 | 73.5 | 20970 | 31.2 | 2214 | 3.30 |
|  | 20<sup>th</sup> May | 112201 | 63336 | 56.4 | 45422 | 40.5 | 3436 | 3.06 |
|  | 30<sup>th</sup> May | 181859 | 89729 | 49.3 | 86934 | 47.8 | 5185 | 2.85 |
|  | 10<sup>th</sup> June | 287157 | 138106 | 48.1 | 140928 | 49.1 | 8108 | 2.82 |
| Maharashtra | 25<sup>th</sup> March | 122 | 122 | 100.0 | 0 | 0.00 | 0 | 0.00 |
|  | 10<sup>th</sup> April | 1574 | 1276 | 81.1 | 188 | 11.94 | 110 | 6.99 |
|  | 20<sup>th</sup> April | 4666 | 3862 | 82.8 | 575 | 12.32 | 232 | 4.97 |
|  | 30<sup>th</sup> April | 10498 | 8266 | 78.7 | 1773 | 16.89 | 459 | 4.37 |
|  | 10<sup>th</sup> May | 22171 | 17140 | 77.3 | 4199 | 18.94 | 832 | 3.75 |
|  | 20<sup>th</sup> May | 39297 | 27589 | 70.2 | 10318 | 26.26 | 1390 | 3.54 |
|  | 30<sup>th</sup> May | 65168 | 34890 | 53.5 | 28081 | 43.09 | 2197 | 3.37 |
|  | 10<sup>th</sup> June | 94041 | 46087 | 49.0 | 44516 | 47.34 | 3438 | 3.66 |
| Tamil Nadu | 25<sup>th</sup> March | 26 | 26 | 100.0 | 0 | 0.00 | 0 | 0.00 |
|  | 10<sup>th</sup> April | 911 | 858 | 94.2 | 44 | 4.83 | 9 | 0.99 |
|  | 20<sup>th</sup> April | 1520 | 1046 | 68.8 | 457 | 30.07 | 17 | 1.12 |
|  | 30<sup>th</sup> April | 2323 | 1038 | 44.7 | 1258 | 54.15 | 27 | 1.16 |
|  | 10<sup>th</sup> May | 7204 | 5198 | 72.2 | 1959 | 27.19 | 47 | 0.65 |
|  | 20<sup>th</sup> May | 13191 | 7221 | 54.7 | 5882 | 44.59 | 88 | 0.67 |
|  | 30<sup>th</sup> May | 21184 | 9021 | 42.6 | 12000 | 56.65 | 163 | 0.77 |
|  | 10<sup>th</sup> June | 36841 | 17182 | 46.6 | 19333 | 52.48 | 326 | 0.88 |
| Delhi | 25<sup>th</sup> March | 35 | 35 | 100.0 | 0 | 0.00 | 0 | 0.00 |
|  | 10<sup>th</sup> April | 903 | 862 | 95.5 | 27 | 2.99 | 14 | 1.55 |
|  | 20<sup>th</sup> April | 2081 | 1603 | 77.0 | 431 | 20.71 | 47 | 2.26 |
|  | 30<sup>th</sup> April | 3515 | 2362 | 67.2 | 1094 | 31.12 | 59 | 1.68 |
|  | 10<sup>th</sup> May | 6923 | 4781 | 69.1 | 2069 | 29.89 | 73 | 1.05 |
|  | 20<sup>th</sup> May | 11088 | 5720 | 51.6 | 5192 | 46.83 | 176 | 1.59 |
|  | 30<sup>th</sup> May | 18549 | 10058 | 54.2 | 8075 | 43.53 | 416 | 2.24 |
|  | 10<sup>th</sup> June | 32810 | 19581 | 59.7 | 12245 | 37.32 | 984 | 3.00 |
| Gujarat | 25<sup>th</sup> March | 38 | 38 | 100.0 | 0 | 0.00 | 0 | 0.00 |
|  | 10<sup>th</sup> April | 378 | 326 | 86.2 | 33 | 8.73 | 19 | 5.03 |
|  | 20<sup>th</sup> April | 1939 | 1737 | 89.6 | 131 | 6.76 | 71 | 3.66 |
|  | 30<sup>th</sup> April | 4395 | 3568 | 81.2 | 613 | 13.95 | 214 | 4.87 |
|  | 10<sup>th</sup> May | 8195 | 5157 | 62.9 | 2545 | 31.06 | 493 | 6.02 |
|  | 20<sup>th</sup> May | 12539 | 6571 | 52.4 | 5219 | 41.62 | 749 | 5.97 |
|  | 30<sup>th</sup> May | 16356 | 6119 | 37.4 | 9230 | 56.43 | 1007 | 6.16 |
|  | 10<sup>th</sup> June | 21554 | 5464 | 25.4 | 14743 | 68.40 | 1347 | 6.25 |
| Rajasthan | 25<sup>th</sup> March | 38 | 38 | 100.0 | 0 | 0.00 | 0 | 0.00 |
|  | 10<sup>th</sup> April | 561 | 498 | 88.8 | 60 | 10.70 | 3 | 0.53 |
|  | 20<sup>th</sup> April | 1576 | 1346 | 85.4 | 205 | 13.01 | 25 | 1.59 |
|  | 30<sup>th</sup> April | 2584 | 1633 | 63.2 | 893 | 34.56 | 58 | 2.24 |
|  | 10<sup>th</sup> May | 3814 | 1465 | 38.4 | 2241 | 58.76 | 108 | 2.83 |
|  | 20<sup>th</sup> May | 6015 | 2464 | 41.0 | 3404 | 56.59 | 147 | 2.44 |
|  | 30<sup>th</sup> May | 8617 | 2685 | 31.2 | 5739 | 66.60 | 193 | 2.24 |
|  | 10<sup>th</sup> June | 11600 | 2772 | 23.9 | 8569 | 73.87 | 259 | 2.23 |
| Uttar Pradesh | 25<sup>th</sup> March | 38 | 38 | 100.0 | 0 | 0.00 | 0 | 0.00 |
|  | 10<sup>th</sup> April | 433 | 397 | 91.7 | 32 | 7.39 | 4 | 0.92 |
|  | 20<sup>th</sup> April | 1184 | 1026 | 86.7 | 140 | 11.82 | 18 | 1.52 |
|  | 30<sup>th</sup> April | 2211 | 1620 | 73.3 | 551 | 24.92 | 40 | 1.81 |
|  | 10<sup>th</sup> May | 3467 | 1735 | 50.0 | 1653 | 47.68 | 79 | 2.28 |
|  | 20<sup>th</sup> May | 5175 | 1982 | 38.3 | 3066 | 59.25 | 127 | 2.45 |
|  | 30<sup>th</sup> May | 7701 | 2837 | 36.8 | 4651 | 60.39 | 213 | 2.77 |
|  | 10<sup>th</sup> June | 11610 | 4318 | 37.2 | 6971 | 60.04 | 321 | 2.76 |

© Source: Covid19india.org

**Figure 1** shows the active cases of India and six selected states. For most of these, active cases have more or less decreased since April 20^th^ with an exception that of Tamil Nadu. Tamil Nadu’s active cases are fluctuating with a sudden increase in the duration April 30^th^ to May 10^th^ and peaking around 10^th^ May. Post 30^th^ May 2020, the active cases line of Uttar Pradesh, Tamil Nadu and Delhi have shown a slight upward trend. Across India, in the said duration, the cases are fluctuating with a small peak around 10^th^ May and then decreasing in the latter part of the taken time period.

**Figure 1:**
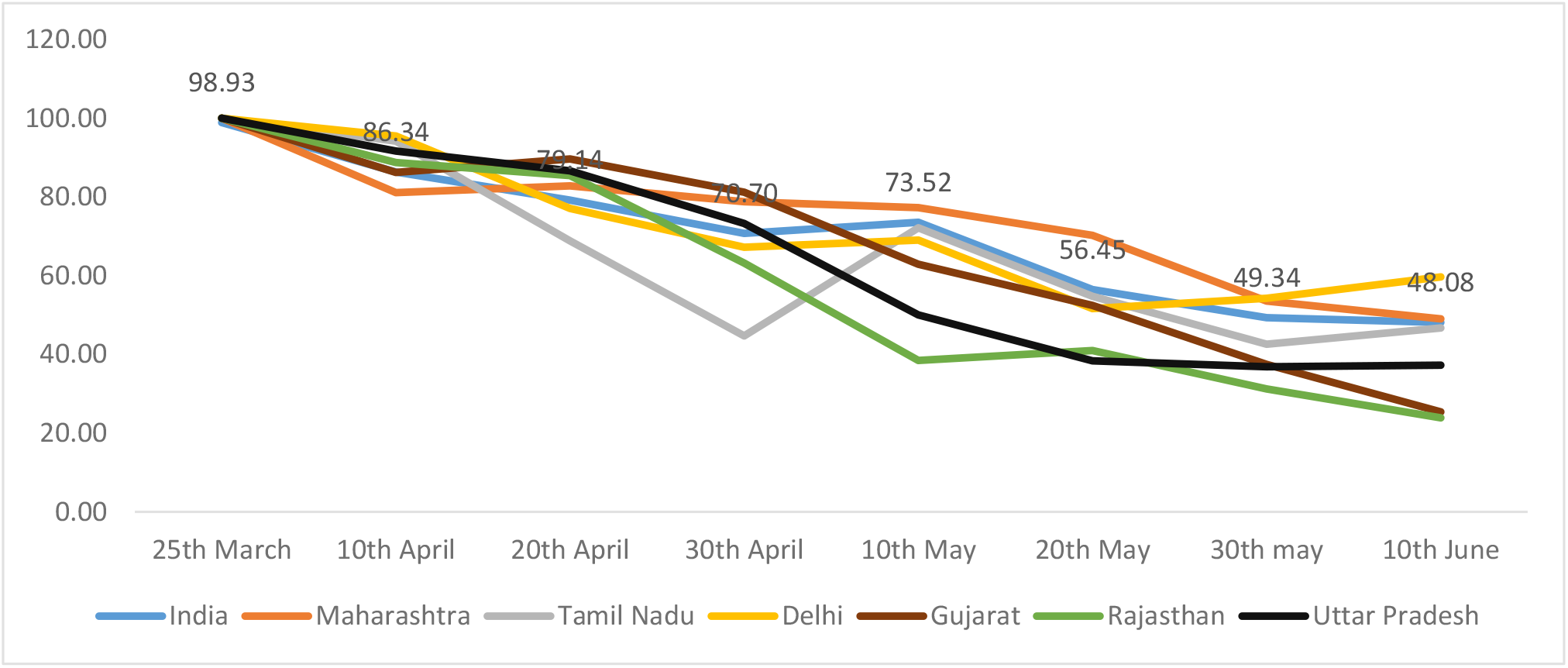
Rate of active cases for India and selected states/UT.

**Figure 2** shows the trend of recovery rates of India and the selected states. The graph shows that the recovery rates are steadily increasing. Tamil Nadu again stands out as the rates can be seen to fluctuate during the period 30^th^ April to 30^th^ May. In this time period, Tamil Nadu sees both high and low peaks in recovery rates. Post 30^th^ May, the recovery rates can be seen to be increasing gradually for India, Maharashtra, Gujarat and Rajasthan. However, a minor decrease can be observed in the recovery rates of Uttar Pradesh, Delhi and Tamil Nadu.

**Figure 2:**
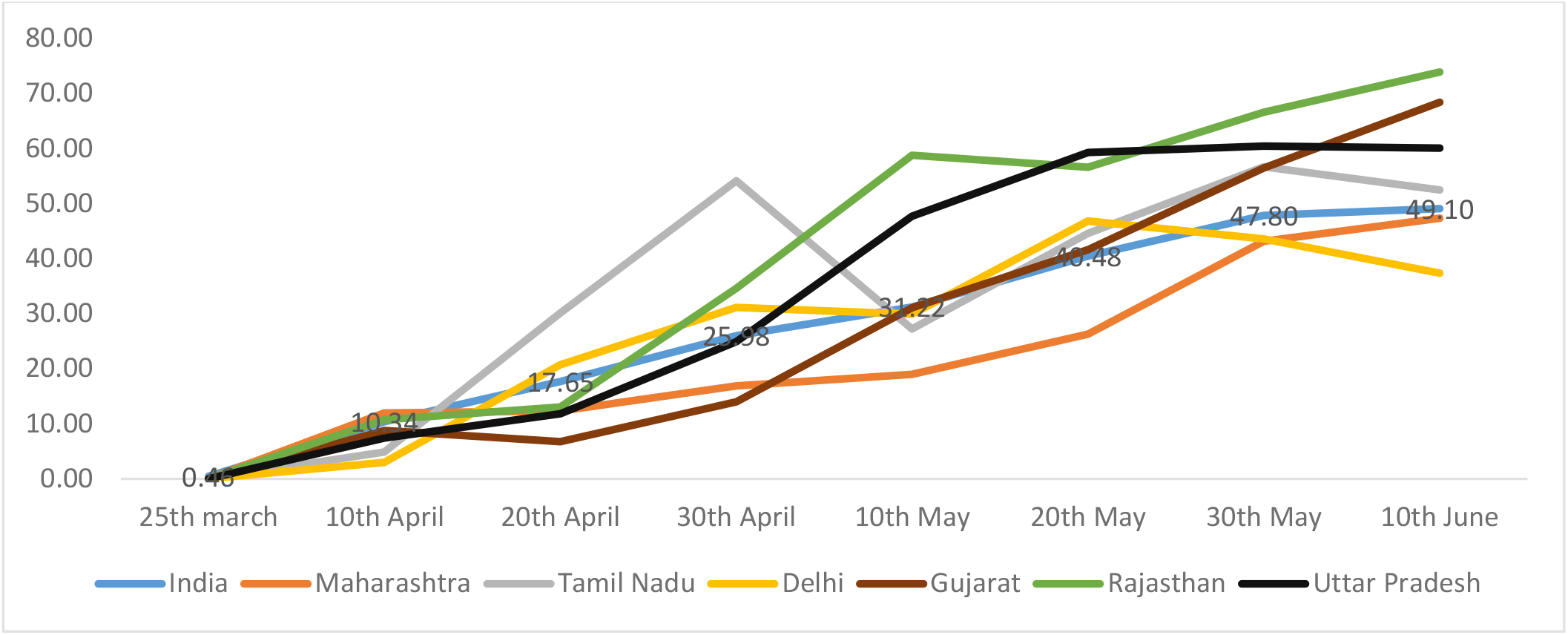
Recovery rate for India and selected states/UT.

**Figure 3** shows the case fatality rate of India and the selected states. It is observed that Tamil Nadu, irrespective of having a higher level of active cases has maintained a very low case fatality rate throughout the duration of analysis. Rajasthan, also, in the entire duration has stayed below the national average of case fatality with Uttar Pradesh rising in the case fatality rates to touch this national average and Delhi surpassing it around 10^th^ June. Maharashtra and Gujarat from the very beginning have been way higher than the national average case fatality rate. However, where case fatality rate of Maharashtra is seen to be decreasing gradually even if still above the national case fatality rate, Gujarat’s case fatality is increasing at an alarming rate. At the end of analysis period, Gujarat’s case fatality rate stands at 6.25 which is way above the national case fatality rate as on 10^th^ June which stood at 2.82.

**Figure 3:**
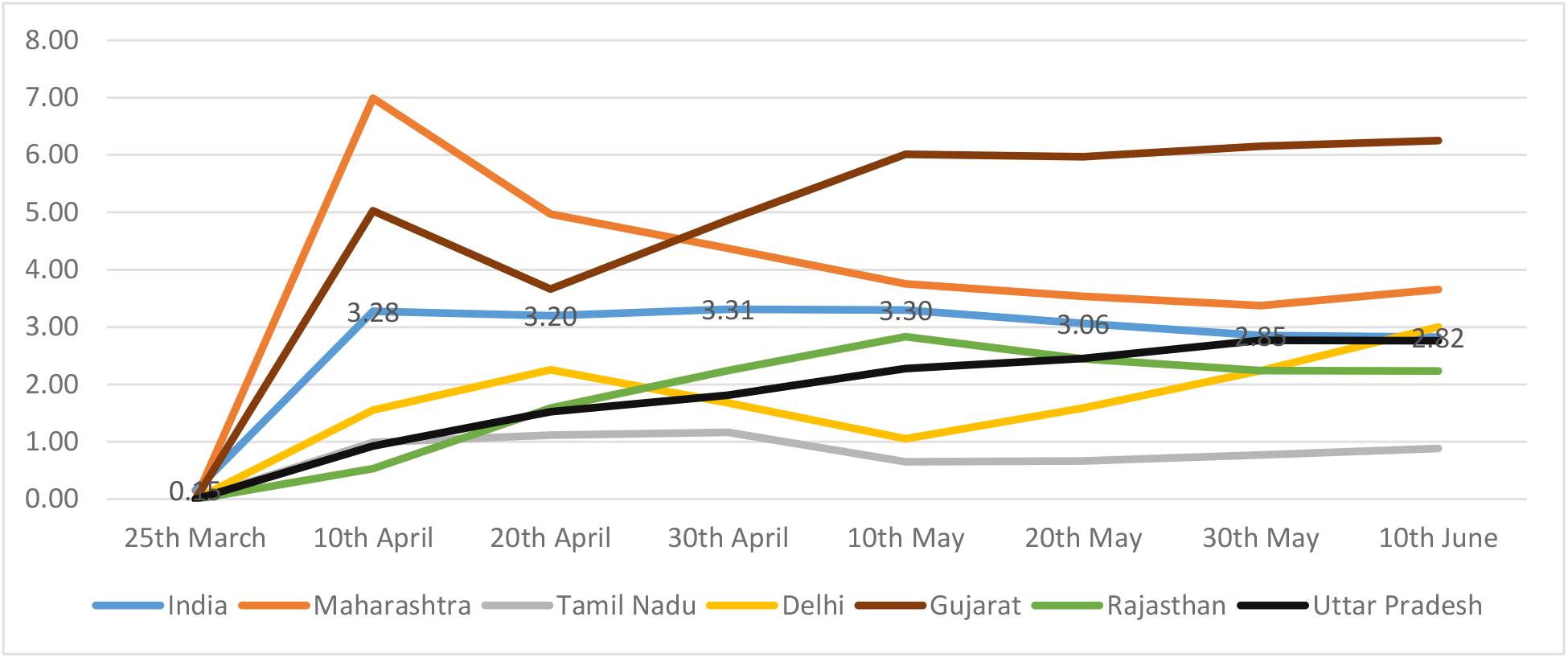
Case fatality rate for India and selected states/UT.

**Table 2** gives the number of test conducted in this duration for India and states/UT and how it increased since the coronavirus reached India. Test positivity rate is calculated and shown in this table and it is defined as proportion of confirmed cases per 100 people tested. The projected population for the year 2020 has been mentioned in the table and it is taken from Report of the technical group on population projections, 2019. This study calculated Tests per million population and cases per million population both have steadily increased. Test per confirmed cases is also calculated and shown in table and it defines the number of test conducted per positive cases.

**Table 2:** **Total test conducted, Confirmed cases, Test positivity rate, Test per million population, Cases per million population, Test per confirmed cases, and Projected Population (2020) for India and selected states/UT for selected time period**.

| Country/States/UT | Date | Total test conducted | Total confirmed cases | Test Positivity rate | Test per million population | Cases per million population | Test per confirmed cases | Projected Population (2020) |
| --- | --- | --- | --- | --- | --- | --- | --- | --- |
| India | 10<sup>th</sup> April | 161330 | 7599 | 4.7 | 120 | 6 | 21.2 | 1347121000 |
|  | 20<sup>th</sup> April | 432104 | 18544 | 4.3 | 321 | 14 | 23.3 |  |
|  | 30<sup>th</sup> April | 830201 | 34867 | 4.2 | 616 | 26 | 23.8 |  |
|  | 10<sup>th</sup> May | 1609037 | 67177 | 4.2 | 1194 | 50 | 24.0 |  |
|  | 20<sup>th</sup> May | 2512388 | 112201 | 4.5 | 1865 | 83 | 22.4 |  |
|  | 30<sup>th</sup> May | 3611599 | 181859 | 5.0 | 2681 | 135 | 19.9 |  |
|  | 10<sup>th</sup> June | 5061332 | 287157 | 5.7 | 3757 | 213 | 17.6 |  |
| Maharashtra | 10<sup>th</sup> April | 30000 | 1574 | 5.2 | 243 | 13 | 19.1 | 123295000 |
|  | 20<sup>th</sup> April | 71321 | 4666 | 6.5 | 578 | 38 | 15.3 |  |
|  | 30<sup>th</sup> April | 135694 | 10498 | 7.7 | 1101 | 85 | 12.9 |  |
|  | 10<sup>th</sup> May | 225524 | 22171 | 9.8 | 1829 | 180 | 10.2 |  |
|  | 20<sup>th</sup> May | 307535 | 39297 | 12.8 | 2494 | 319 | 7.8 |  |
|  | 30<sup>th</sup> May | 448661 | 65168 | 14.5 | 3639 | 529 | 6.9 |  |
|  | 10<sup>th</sup> June | 595282 | 94041 | 15.8 | 4828 | 763 | 6.3 |  |
| Tamil Nadu | 10<sup>th</sup> April | 8410 | 911 | 10.8 | 111 | 12 | 9.2 | 76049000 |
|  | 20<sup>th</sup> April | 46985 | 1520 | 3.2 | 618 | 20 | 30.9 |  |
|  | 30<sup>th</sup> April | 119748 | 2323 | 1.9 | 1575 | 31 | 51.5 |  |
|  | 10<sup>th</sup> May | 243037 | 7204 | 3.0 | 3196 | 95 | 33.7 |  |
|  | 20<sup>th</sup> May | 360068 | 13191 | 3.7 | 4735 | 173 | 27.3 |  |
|  | 30<sup>th</sup> May | 479155 | 21184 | 4.4 | 6301 | 279 | 22.6 |  |
|  | 10<sup>th</sup> June | 638846 | 36841 | 5.8 | 8400 | 484 | 17.3 |  |
| Delhi | 10<sup>th</sup> April | 11061 | 903 | 8.2 | 548 | 45 | 12.2 | 20193000 |
|  | 20<sup>th</sup> April | 25900 | 2081 | 8.0 | 1283 | 103 | 12.4 |  |
|  | 30<sup>th</sup> April | 47225 | 3515 | 7.4 | 2339 | 174 | 13.4 |  |
|  | 10<sup>th</sup> May | 93810 | 6923 | 7.4 | 4646 | 343 | 13.6 |  |
|  | 20<sup>th</sup> May | 150282 | 11088 | 7.4 | 7442 | 549 | 13.6 |  |
|  | 30<sup>th</sup> May | 206739 | 18549 | 9.0 | 10238 | 919 | 11.1 |  |
|  | 10<sup>th</sup> June | 266156 | 32810 | 12.3 | 13181 | 1625 | 8.1 |  |
| Gujarat | 10<sup>th</sup> April | 7718 | 378 | 4.9 | 112 | 5 | 20.4 | 68862000 |
|  | 20<sup>th</sup> April | 33316 | 1939 | 5.8 | 484 | 28 | 17.2 |  |
|  | 30<sup>th</sup> April | 64007 | 4395 | 6.9 | 929 | 64 | 14.6 |  |
|  | 10<sup>th</sup> May | 113493 | 8195 | 7.2 | 1648 | 119 | 13.8 |  |
|  | 20<sup>th</sup> May | 160772 | 12539 | 7.8 | 2335 | 182 | 12.8 |  |
|  | 30<sup>th</sup> May | 205780 | 16356 | 7.9 | 2988 | 238 | 12.6 |  |
|  | 10<sup>th</sup> June | 266404 | 21554 | 8.1 | 3869 | 313 | 12.4 |  |
| Rajasthan | 10<sup>th</sup> April | 22349 | 561 | 2.5 | 286 | 7 | 39.8 | 78273000 |
|  | 20<sup>th</sup> April | 57290 | 1576 | 2.8 | 732 | 20 | 36.4 |  |
|  | 30<sup>th</sup> April | 103704 | 2584 | 2.5 | 1325 | 33 | 40.1 |  |
|  | 10<sup>th</sup> May | 166424 | 3814 | 2.3 | 2126 | 49 | 43.6 |  |
|  | 20<sup>th</sup> May | 265555 | 6015 | 2.3 | 3393 | 77 | 44.1 |  |
|  | 30<sup>th</sup> May | 395490 | 8617 | 2.2 | 5053 | 110 | 45.9 |  |
|  | 10<sup>th</sup> June | 543312 | 11600 | 2.1 | 6941 | 148 | 46.8 |  |
| Uttar Pradesh | 10<sup>th</sup> April | 9332 | 433 | 4.6 | 41 | 2 | 21.6 | 227943000 |
|  | 20<sup>th</sup> April | 34326 | 1184 | 3.4 | 151 | 5 | 29.0 |  |
|  | 30<sup>th</sup> April | 78013 | 2211 | 2.8 | 342 | 10 | 35.3 |  |
|  | 10<sup>th</sup> May | 129955 | 3467 | 2.7 | 570 | 15 | 37.5 |  |
|  | 20<sup>th</sup> May | 191164 | 5175 | 2.7 | 839 | 23 | 36.9 |  |
|  | 30<sup>th</sup> May | 279288 | 7701 | 2.8 | 1225 | 34 | 36.3 |  |
|  | 10<sup>th</sup> June | 404637 | 11610 | 2.9 | 1775 | 51 | 34.9 |  |

© Source: Covid19india.org

**Table 3:** **List of Covid-19 designated government and private laboratories provided by ICMR on June 10**^**th**^ **2020 for India and selected states/UT**.

| States/UTs | Total Operational Laboratories | Government Laboratories | Private Laboratories |
| --- | --- | --- | --- |
| Maharashtra | 95 | 53 | 42 |
| Tamil Nadu | 77 | 44 | 33 |
| Delhi | 40 | 17 | 23 |
| Gujarat | 41 | 23 | 18 |
| Rajasthan | 24 | 21 | 3 |
| Uttar Pradesh | 75 | 66 | 9 |
| **India** | **837** | **602** | **235** |

© Source: Covid19india.org

**Figure 4** shows the test positivity rate means the proportion of confirmed cases per 100 people tested. India’s test positivity rate has increased from 4.7 to 5.6 showing steady growth. Gujarat has shown rise in curve in initial phase till April but now is almost steady. The curve of Maharashtra and Delhi is showing upward trend, with the number of cases increasing on daily basis the test positivity rate of these states are increasing. Rajasthan and Uttar Pradesh are at lower level since beginning.

**Figure 4:**
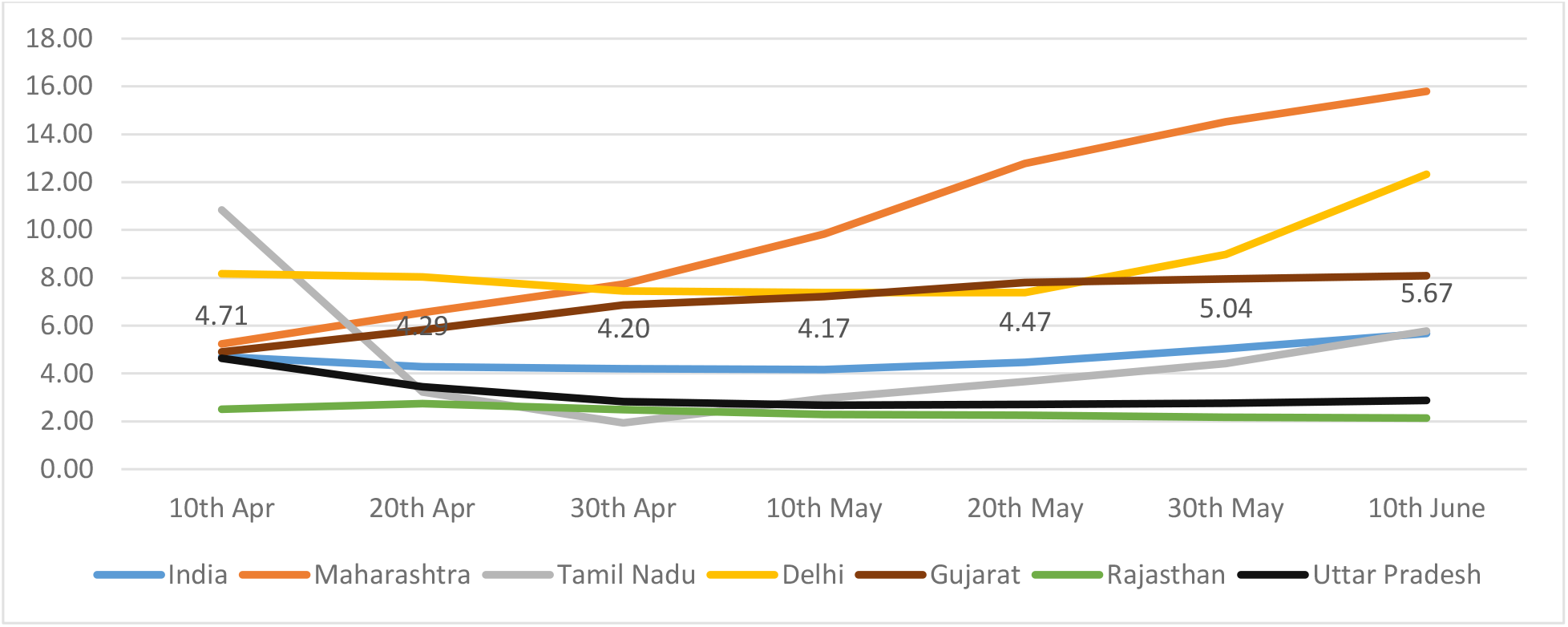
Test positivity rates for India and selected states/UT.

**Figure 5** shows tests conducted per million population. As it can be seen, testing per million population has steadily increased during the analysis duration. Although, the over-all India level testing remains at a low of 3757 tests per million compared to other affected countries. Delhi can be seen to be flaring well as far as tests are concerned. As on 10^th^ June, Delhi’s stood at 13181 tests per million population. Delhi is followed by Tamil Nadu and Rajasthan. Maharashtra although with higher number of active cases, still seems to be lagging behind in testing.

**Figure 5:**
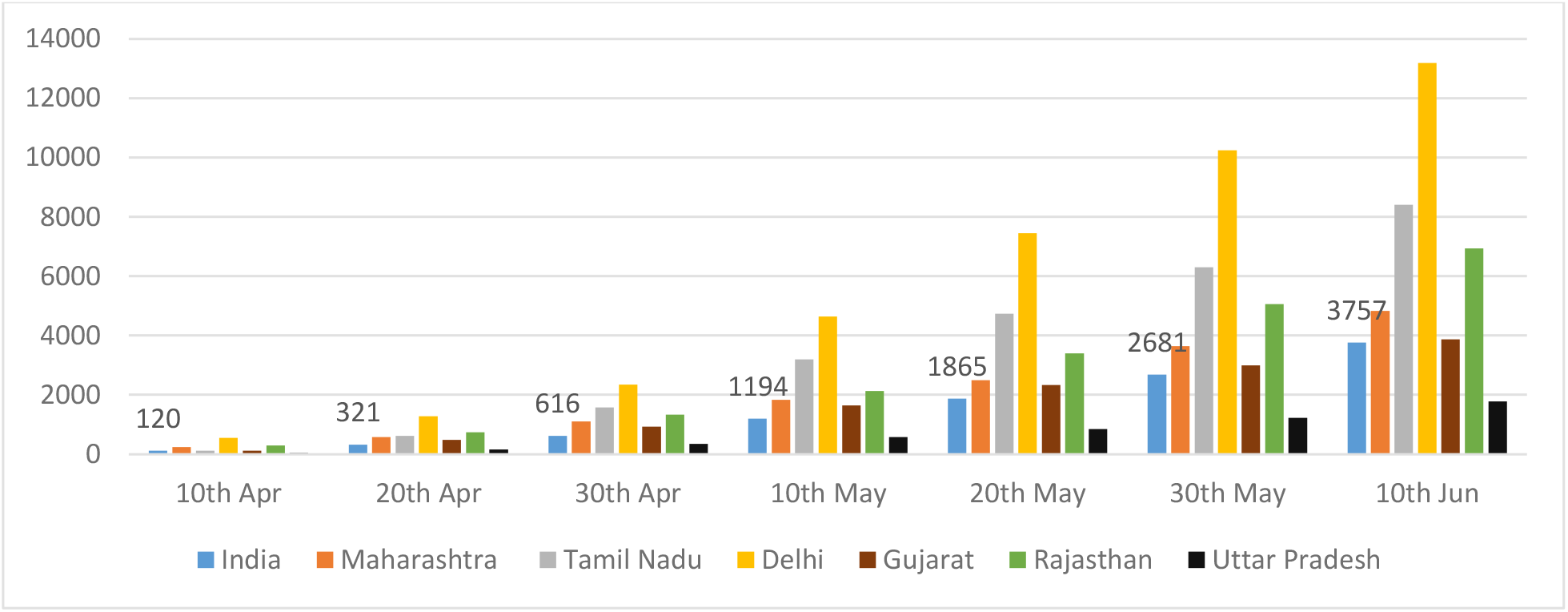
Testing per million population (projected 2020) for India and selected states/UT.

**Figure 6** shows cases per million population across India. Delhi has shown continuous increase in cases per million population and has reached 1600 mark followed by Maharashtra but still a wide gap exists between these two states with Maharashtra at almost half of Delhi. States like Tamil Nadu and Gujarat were growing but now have shown a considerable rise after announcement of unlock 1.0. Apart from these states, Rajasthan and Uttar Pradesh are showing low cases per million population compared to other states since beginning.

**Figure 6:**
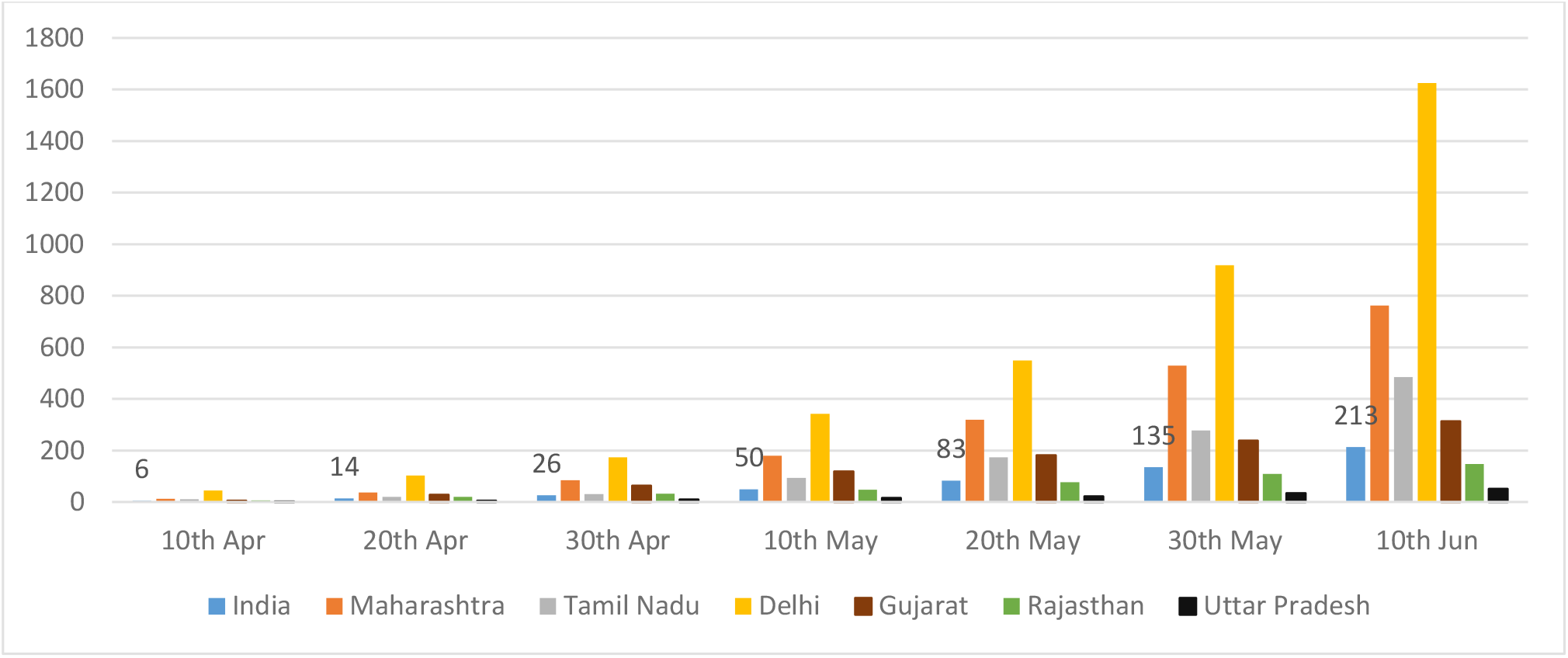
Cases per million population (as projected for 2020) of India and selected states/UT.

**Figure 7** shows tests per confirmed cases. As the curve declines the severity of confirmed cases is increasing. As on June 10^th^ India’s curve is showing little decline as the lockdown lifted up. Severity is high in Maharashtra as every 6th person who is tested is coronavirus positive followed by Delhi as every 8^th^ person is coronavirus positive. Whereas states which are showing better results are Uttar Pradesh where every 35^th^ is coronavirus positive and in Rajasthan every 47^th^ person is coronavirus positive.

**Figure 7:**
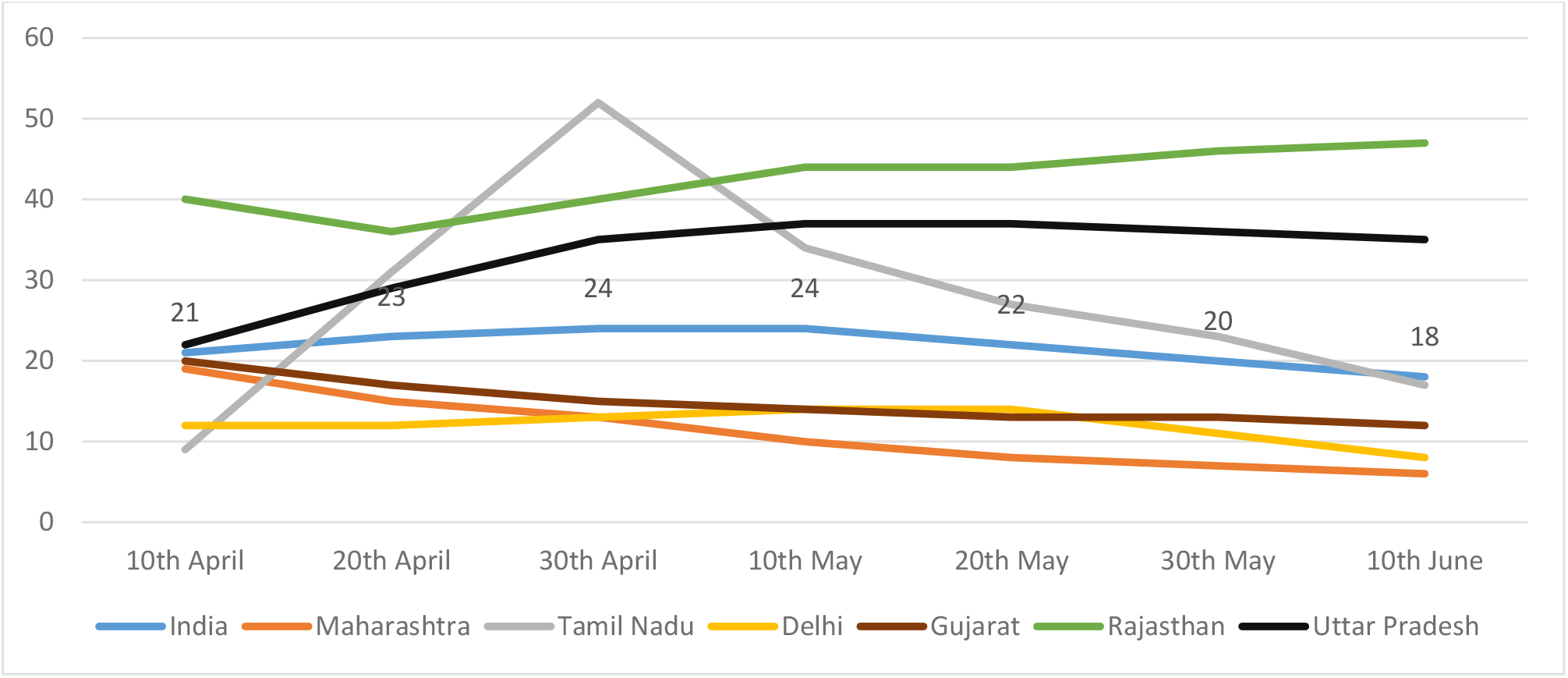
Tests per confirmed cases for India and selected states/UT.

**Figure 8** shows the percentage of government and private laboratories doing Covid-19 tests. More the inclusion of private laboratories for Covid-19 tests shows that state is increasing its efficiency of testing. Around 28 percent of Covid-19 designated laboratories are private in India. In states like Maharashtra, Tamil Nadu, and Gujarat more than 40 percent of laboratories are private whereas in Delhi more than half of the laboratories are private. In Uttar Pradesh and Rajasthan number of private laboratories doing Covid-19 tests is around 12 percent.

**Figure 8:**
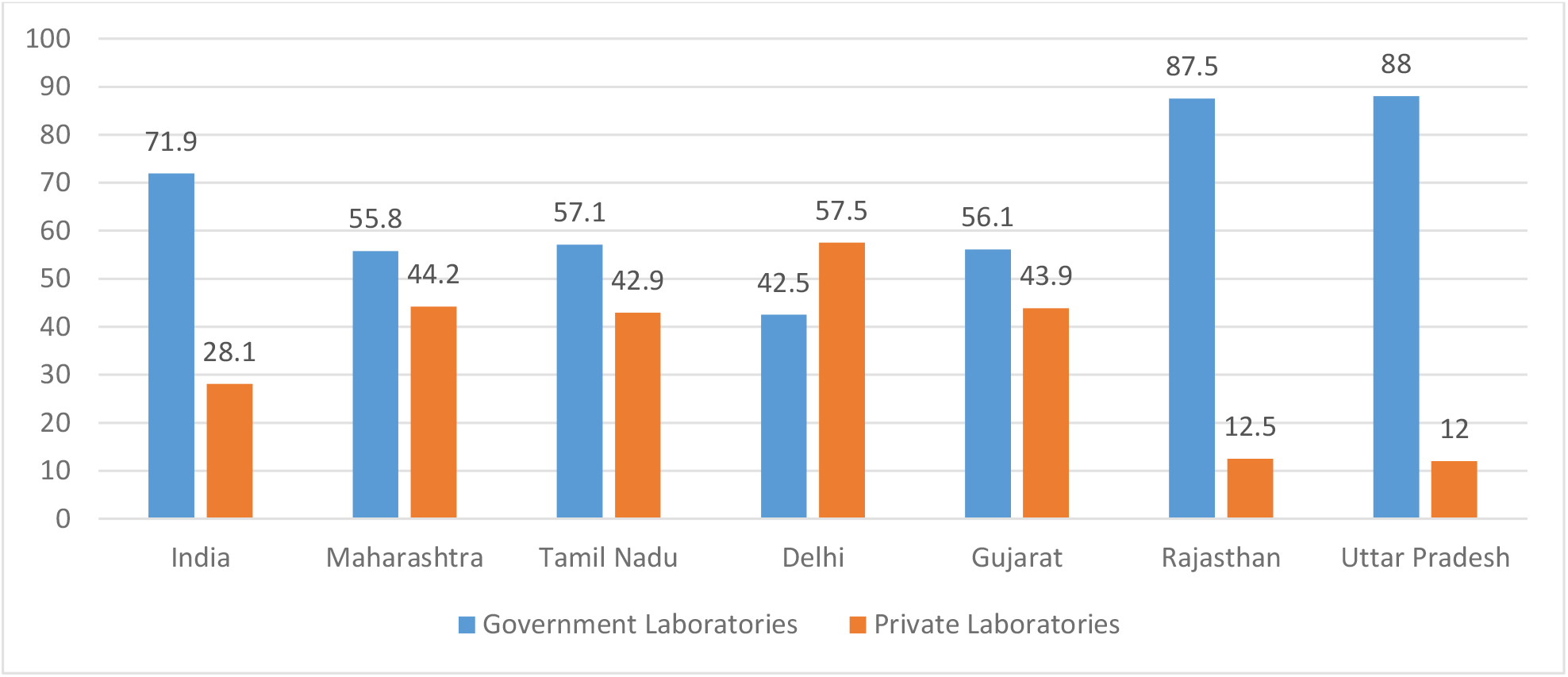
Percentage of Covid-19 designated government and private laboratories as per ICMR on June 10^th^ 2020, for India and selected states/UT.

## Discussion and Conclusion

Since the outbreak of novel coronavirus, Government of India took a series quick actions and as a part of it announced a nationwide lockdown on March 25^th^ 2020. Measures like social distancing, covering face and nose, maintaining hygiene and avoiding public gathering were made compulsory. The measures were to ensure the break of human chain for transmission and thus ultimately tackle the SARS CoV-2 spread. The government has been trying to beat Covid-19 at two fronts, one, flatten the curve of coronavirus positive cases and second, to restrict the exponential growth in the number of positive cases ^[14]^. However with the announcement of first unlock on June 1^st^ 2020, the statistics are bound to change.

This study, has thus, tried to show how India’s numbers have behaved since lockdown to till June 10^th^ i.e. 10 days into the unlocked India and the strategies used by Government of India to counter Covid-19.

The Ministry of Health and Family Welfare stated on June 12^th^ that India’s doubling rate of Covid-19 cases has increased from 3.4 days when lockdown began (March 25^th^) to 17.4 days currently^[15]^. Along with it India’s active cases rate and case fatality rate are decreasing whereas recovery rate is showing an upward trend since the outbreak. At present, recovery rate is higher than active cases rate, however this is not the point to let our guards down as the peak is yet to reach of the pandemic. Also, recovery rates talks about cumulative/accumulated numbers whereas active rate give the current situation thus making them non-comparable ^[16]^. Simultaneously, an analysis of the current situation in terms of new active cases and new recovered cases (in the duration of 10 days from unlock 1.0), the situation is opposite, the active cases are more than recovered cases ^[17]^.

This study highlighted the performance of six states but the states that stood out were Maharashtra, Tamil Nadu, and most importantly Delhi, which were well below assuring level as there active cases are not only fluctuating but also increasing as India’s move to unlock 1.0. Number of new cases coming on a daily basis are alarming in these major states making them a contributor to more than 70 percent Covid-19. Test positivity rate clearly show how the cases are accelerating with Maharashtra and Delhi as the top contributors.

However, test per million population of India and in the selected states are increasing. An increase in number of testing can trace an infected person at an earlier stage and thus inhibit its transmission at the same time since its source would be recognizable, it’ll save the states from reaching stage three of CoV-2 transmission. Indian Council of Medical Research has granted private laboratories the access to conduct testing for Covid-19 and hence this has led to a better and increased capacity and efficiency of testing. With the increase in testing the cases per million population has increased and this comes out as a pressing matter of concern for India. The study has highlighted the situation of Delhi and Maharashtra that have become major hotspots as test per positive cases in Maharashtra is 6 percent and in Delhi it is 8 percent which means in Maharashtra out of every 6 person tested one is coming as coronavirus positive and in Delhi out of every 8 persons tested one is coming positive. This graph is going down steadily for all the selected states and for India as whole meaning that the spread is once again gaining pace.

As the daily cases are increasing the government has also realized the need of more hospitals in India. Therefore, the number of Covid-19 designated hospitals have been increased by 22 percent i.e. from 2719 Covid-19 Health Centers on May 7^th^ to 3319 Covid-19 Health Centers on June 5^th [18]^. With the inclusion of private labs and increasing number of health centers government is trying to counter the Covid-19 crisis with increased testing and accessibility of hospitals for infected patients, but even after this we are grappling to match pace with the increasing Covid-19 cases especially in Delhi and Maharashtra. The number of active and newly infected cases are showing a tremendous increase in these 2 states.

India has spent only 1.28 percent of GDP on its health infrastructure which is quite less when compared to other developed nation where pandemic is at peak. As per Global health expert’s prediction India lags in health infrastructure and capability to deal with this pandemic ^[19]^. Though India has performed well in wake of the SARS CoV pandemic, it can be credited to timely lockdown thereby actively inhibiting its transmission yet it struggled to flatten the curve of infected patients.

However lockdown has delayed the peak of coronavirus in Indian subcontinent giving it time to strengthen its health infrastructure and thus be better equipped to fight the upcoming challenge. Government of India is trying to tackle the pandemic with joining hands and involving both public as well as private laboratories and hospitals to share the burden. However, to ensure the affordability along with accessibility government needs to have an upper ceiling of the charges that people would have to pay in order to get treated.

Thus it can be said that India fared well at the beginning with timely lockdown and series of quick actions but this was at cost of economy. With new regime of unlock 1.0, India is reluctant to ignore economy anymore and thus India’s initial success of containing the spread of Cov-2 seems to be faltering. India is trying to counter the pandemic with public-private partnership as well as an increase in efficiency and accessibility of testing and health care facility but the affordability still remains an aspect that can be worked upon. Where on one hand, India is grappling to take control of the situation with increase in daily positivity rates and tests per positive cases, on the other hand, if experts are to be believed, we are yet to see the worst of it in around mid-July to August when Cov-2 would reach its actual peak. Hence, it goes without saying that it isn’t the end of pandemic and even though lockdown has been lifted, it’d be advisable to not venture out unnecessarily and maintain social distancing, avoiding public places and covering mouth and nose along with the recommended hygiene practices.

## Data Availability

India Covid-19 tracker 2020.

## Notes

### Competing Interest Statement

The authors have declared no competing interest.

### Funding Statement

No funding received.

